# Urinary Extracellular vesicles abundance of SLC12A3 (NCC) increase and Aquaporine2 decrease following DASH diet implementation

**DOI:** 10.1101/2022.11.29.22282878

**Authors:** Dana Bielopolski, Luca Musante, Henrik Molina, Douglas Barrows, Thomas Carrol, Jonathan N. Tobin, Rhonda. G Kost, U Erdbruegger

## Abstract

The Dietary Approach to Stop Hypertension (DASH) diet is a proven intervention to treat hypertension, yet its mechanism is not clearly known. We investigated the change in protein abundance patterns in urine extracellular vesicles (uEV’s) following DASH diet implementation.

A pilot study was carried out to compare uEVs isolated using three different methods: a low centrifugation (P20), high centrifugation (P100), and a combination of both (P20 and P100). Uromodulin was removed by size exclusion chromatography and low ionic strength washing. Mass spectrometry analysis identified 1,593 proteins in the combined fraction (P20+P100), 1434 in the P20 fraction and 1229 in the P100 fraction. The combined fraction was chosen for further analysis. Statistical analysis was carried out using R and Limma to identify all proteins that changed before and during 11 days of DASH intervention (p < 0.05) as well as between individual timepoints.

Nine hypertensive volunteers were admitted for a 14-day supervised transition from American style diet to DASH diet. First-void urine was collected on days 0, 5, 11 for uEV processing. In total, 1800 proteins were identified across all 27 DASH samples with 22 proteins upregulated and 25 down regulated between day 0 and both days 5 and 11. These included increased abundance of SLC12A3 (NCC) and reduced abundance of Aquaporine 2.

These changes could explain the increased urine volume and reduced sodium reabsorption that lead to blood pressure reduction following consumption of the high potassium and low sodium DASH diet. uEVs may serve as a surrogate to a more invasive procedure.

**Translational Statement:** Myriad studies have characterized blood pressure reduction following DASH diet implementation, yet its precise mechanism is unclear. Here, we demonstrate for the first time the effect of DASH nutritional changes, on kidney ion channel composition using proteomic analysis of urinary EVs. Using this innovative tool as a substitute for an invasive procedure, we show that the expression of Aquaporine2 increases and NCC decreases in response to DASH which may account for its antihypertensive effect. Our results indicate that urinary EV are a potential biomarker for DASH compliance, and targeting aquaporine2 may be an effective innovative therapeutic strategy for blood pressure reduction.

## Introduction

Hypertension is a disease of the Western world as it is worsened by sedentary life style, and can be controlled by low salt diet, weight loss and smoking secession^1^. Salt sensitive hypertension is found in over 50% of cases and is defined as blood pressure (BP) that exhibits changes parallel to changes in salt intake^2^. It is commonly accepted that in response to increased salt intake, salt excretion can increase up to a certain threshold, after which salt and water will accumulate, serum volume increases and consequently blood pressure increases^3^.

Hypertension and salt sensitivity can be attenuated by increaseing potassium intake as documented in numerous epidemiological and interventional studies. ^4^ The dietary approach to stop hypertension (DASH) diet, a low salt but high potassium diet, is a recommended intervention to lower blood pressure and is more effective for every sodium level compared to low salt diet^5^ alone. Mounting evidence suggests that the potassium component of the diet acts as a switch in the distal convoluted tubule^6^ to reduce sodium reabsorption, similar to a diuretic but without the side effects.

Several trials have attempted to elucidate the antihypertensive effect of DASH diet, yet none have reached a definitive conclusion. ^7–11^ One proposed mechanism is an increase in nitric oxide bioavailability, as measured by plasma nitrite following a stressor, with possible subsequent effects on vascular basal tone^11^. It was also suggested that phenols and their derivatives lower BP by scavenging reactive oxygen species in the vasculature and inhibiting vascular smooth muscle cell proliferation^12^. Other studies have focused on augmented furosemide-sensitive Na+-K+-2Cl2 cotransporter activity^13^, and an increased ratio of long WNK1 kinase to kidney-specific WNK1, resulting in renal Na+ retention and salt-sensitive hypertension in those with low dietary K+ intake^14^. Since these trials were based on animal models, none were able to characterize changes in human ion channel protein abundance following nutritional changes, and avoid invasive procedures.

Urinary extracellular vesicles (uEVs) are nanosized vesicles released from epithelial cells within the kidney and the urinary tract. uEVs hold the promise of use as non-invasive read-out of kidney cellular function in health and disease. Their protein cargo has been shown to mirror tissue content^15–17^ and hence might serve as a surrogate marker for physiologic and pathophysiologic changes occurring within the kidney. To examine the underline mechanism of the DASH diet in adults with mildly elevated blood pressure who were not taking any antihypertensive medications, we conducted an inpatient open label nutritional study transitioning hypertensive volunteers from an American style diet to DASH diet^18^.We witnessed an increase in serum aldosterone five days into the intervention, paralleling a reversal in urinary electrolyte ratio of sodium to potassium. Urine collected from our participants was then used to characterize changes in uEV protein abundance in order to understand non-invasively changes occurring in the kidney epithelium in response to DASH.

## Methods

<u>Patients, dietary intervention, and sample collection</u> were previously described^18^. Briefly, 9 volunteers, with stage I or stage II hypertension but otherwise healthy, were admitted for a 2 week inpatient stay at the Rockefeller University Hospital. They transitioned from American style diet to DASH diet, consuming menus meticulously designed by a bionutrionist to include the appropriate amount of electrolytes^19^. Urine samples were collected daily, in a container with a protease inhibitor (cOmplete; Roche, Woerden, The Netherlands), one tablet per 10 ml urine and centrifuged (3000*g* for 5 minutes) to remove cells and debris before storage at −80°C until processing.

### uEV separation for assessment of uEV protein analysis with Mass spectrometry (a detailed description can be found in the supplementary material)

For this proof of concept study we generated uEV preparations as triplicates without contamination with uromodulin and compared uEVs enriched with three methods: a low centrifugation (P20), a high centrifugation force (P100) and a combination of both (adding P20 to P100) ^20^. A majority of researchers focus on analysis of the P100 uEV pellet, however our preliminary work showed that the low centrifugation pellet P20 is a very valuable source of uEVs and should not be discarded. In brief, one healthy volunteer collected first morning void urine (150ml) with approval from our institutional ethic board (IRB). This sample was processed in three replicates (triplicates). Urine was processed within 3 hours without adding any protease inhibitors.

Urine sample (15ml) was centrifuged at 4600g for 30 minutes to remove cell debris. The pellet was discarded, and the supernatant was then centrifuged at 21,200g (further details of the centrifugation method are summarized in the supplementary material). The first resulting pellet, termed P20, was used as the low centrifugation pellet for Mass Spectrometry analysis, while the supernatant was further centrifuged at 164,244g. The second resulting pellet was termed P100. Both pellets were further processed to remove Uromodulin. In particular, P20 pellet was treated with low ionic strength buffer (10 mM HEPES pH 7.4) supplemented with EDTA (2.5 mM pH 8.0) to depolymerize Uromodulin^20,21,22^. Chloroform and methanol were used for de-lipidation to precipitate this P20 pellet for proteomic analysis. uEV pellets after removal of bulk of Uromodulin of P20 and P100 are visualized with PAGE separation followed by total protein staining with Coomassie (see Figure 2 C and E). The removal of Uromodulin (or Tamm-Horsfall protein, THP) from the P100 pellet was accomplished by using size exclusion chromatography. The uEV P100 pellet was resolubilized in guanidine 6M in acidic condition (PH=3.5) and mixed for 20 minutes in end-over-end wheel before being applied on top of size exclusion chromatography (SEC) 70nm original column (IZON). Columns were washed with 2 mL guanidine buffer before samples loading and 1mL after loading to facilitate an unfolding environment allowing the separation of Uromodulin from uEVs. Completion of chromatography was evident by elution of blue bromophenol dye. The collection of the fraction was carried out with the aid of automatic fraction collector. One mL fraction was collected and precipitated by Trichoroacetic Acid and Sodium Deoxycholate (TCA-DOC) Presence of uEV’s was validated by Western blotting for TSG101 (see Figure 2 D and F). Chloroform methanol and acetone were used to precipitate the pellet for proteomic analysis. In addition to preparing the P20 and P100 pellet for Mass Spectrometry we also combined the P20 and P100 (after uromodulin removal) and performed also a Chloroform methanol and acetone extraction to precipitate the combined uEV pellet.

**Figure 1:**
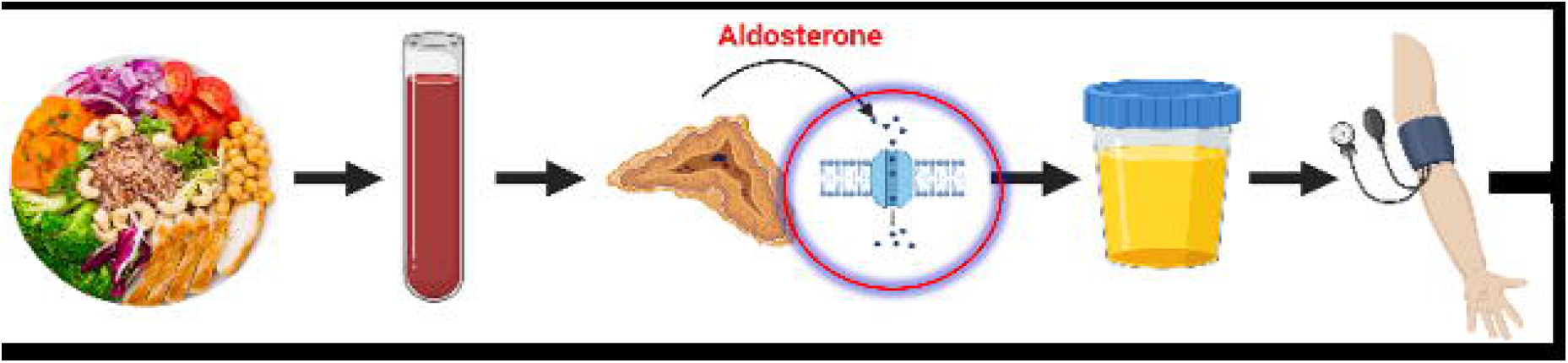
Model depicting the proposed mechanism of DASH diet antihypertensive effect. Cascade of events from implementation of nutritional changes to serum level of electrolytes, secretion of aldosterone resulting in changed abundance pattern of kidney ion channels. Following that change there is a change in urine electrolytes ratio, blood pressure reduction and eventually improved cardiovascular health.

**Figure 2:**
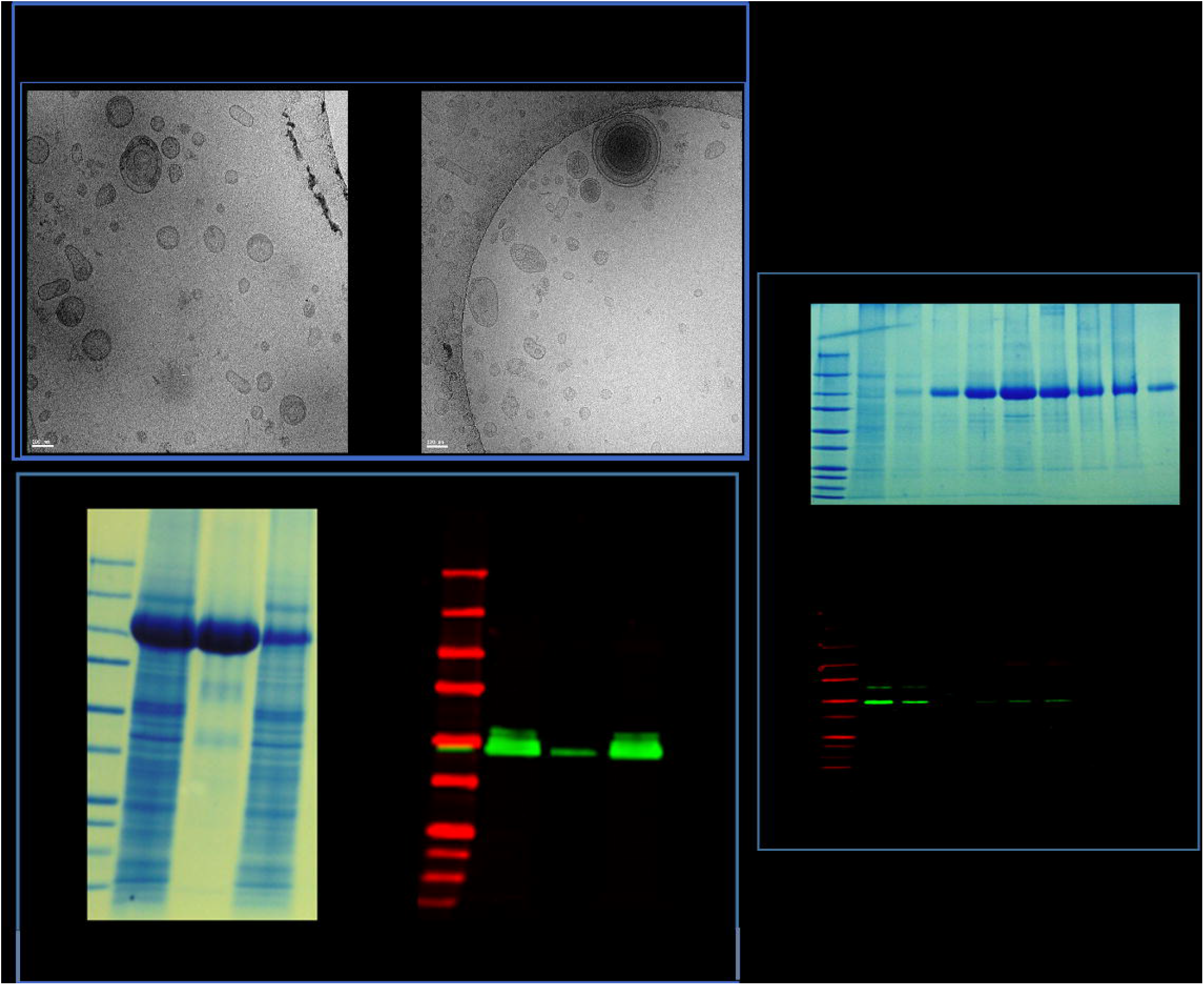
uEV extraction validation from the different fractions. Panels A, B show cryo-TEM of EV’s from P20 treated with low ionic strength buffer, and the 1^st^ two fractions of P100. Panels C, E are Coomassie blue stained gels showing a bulk of protein at a height of 100 (y axis) corresponding to uromodulin. Panel C relates to P20, untreated (1), the supernatant treated with low ionic strength buffer (2) and the pellet after treatment(3). Panel E corresponds to P100 fractionation by SEC (x axis). Panels D F are western blot analysis of the membrane corresponding to the gel, reacted with antibody directed against TSG101. SEC – size exclusion chromatography.

### uEV separation from urine of DASH intervention study

first void spot urine samples collected on days 0, 5 and 11 (volume 20 ml) were thawed and recentrifuged. Fifteen mL of urine were used to enrich urinary EVs by differential centrifugation to generate a low (20,000G, P20) and high (100,000G, P100) centrifugation EV pellet (latter from supernatant of initial 20,000 G CF). Uromodulin (THP) was removed from both pellets, as described in the supplementary material by low ionic strength buffer (P20) and SCE (P100) as described for the pilot study samples. The P100 pellet was resolubilized in 6 M guanidine in acidic condition and applied to a size exclusion chromatography (70nm) (Supplementary Figure 2). Both uEV pellets (form P20 and P100) were combined for further analysis according to the results from our proof-of-concept study as outlined above and discussed in results and discussion below. Protein pattern was analyzed by Western blotting for TSG101 detection (Sigma).

### Size analysis with Nano size Tracking Analysis (NTA) (Supplementary Table 6)

The analysis was performed with Zetaview (Particle Metrix) instrument equipped with 520 nm laser, 550 nm long pass cut off filter and sCMOS camera. DI waters were filtered on the day of analysis through 0.22 μm syringe filter.

uEV labeling was done using Exoglow fluorescent NTA labeling kit from System Biosciences according to manufacturer’s protocol. Briefly, 12 μL of reaction buffer were mixed with 2 μL of dye and 14 μL of sample. The mixture was vortexed for 15 seconds to mix well and samples were incubated at RT for 10 minutes. Liposomes (provided with kit) were used as labeling control: 1 μL of liposomes was mixed with 12 μL of reaction buffer and 2 μL of dye. Dilutions were made by mixing DI water filtered through 0.2 μm syringe filter with corresponding volume of a sample.

### Proteomic analysis

Mass spectrometry of samples enriched for EVs was performed at The Rockefeller University Proteomics Resource Center, as described^23^. Briefly, proteins were precipitated and re-dissolved in 50uL 8M Urea/50mM ammonium bicarbonate/20mM Dithiothreitol followed by alkylation (iodoacetamide). Proteins were first digested with Endoproteinase Lys-C at less than 4M Urea and then diluted to less than 2M Urea for trypsinization. Digestions were halted by adding neat trifluoroacetic acid (TFA) and peptides were subjected to Solid Phase Extraction (C18-like Empore). Peptides were separated using a 12cm packed-in-emitter column (inner diameter of 75um) and data were acquired on an Q-Exactive HF mass spectrometer (Thermo Fisher Scientific). Data was searched against a Uniprots Human database, using Proteome/Discoverer+Mascot as well as MaxQuant.(version 2.0.3.0) using a False Discovery Rate of 1% for proteins and 2% for peptides. For the pilot study only proteins quantified in at least two of three replicates in at least one group were retained, GENE-E software was used for heatmap generation and data display^24^.

### Statistical analyses

were performed using R. Log transformed IBAQ values were quantile normalized using the Bioconductor Limma package^25^ (version 3.50.1) following imputation of missing values using the fitdistr function within the MASS R package^26^ (version 7.3-54). The toptableF and topTable functions implemented within Limma were used to identify all proteins detected by mass spectrometry that changed across the time course (p < 0.05) as well as between individual timepoints. Adjustment for patient effects for visualization was performed using the removeBatchEffect function in Limma.

The abundance of differential proteins was visualized using the heatmap R package (1.0.12) with a heatmap showing the z-scores of each protein across all samples (row scaled). Z-score is calculated as (value – row mean)/(row standard deviation).

Gene Ontology (GO) analysis was performed using DAVID annotation tool^27^ with uEV proteins changing throughout the time course. Input was the combination of uEV enriched proteins (iBAQ count in uEV on days 0,5, 11, Log2 >0), and the background was all identified proteins in the pool (defined by at least one iBAQ value). We uploaded protein lists that changed expression from day 0 to days 5 and 11. GO terms with enrichment fold >1, passing the false discovery rate 5% threshold were retained. GO functional analysis was performed based on log2 fold-change in uEV >1 or less than −1. KEGG was run under the Cytoscape framework (v3.8.2), with GO Biological Process, Molecular Function, Cellular Component (all annotations updated on February 11, 2021), and (annotation updated on February 8, 2021) in use. Evidence level was set at “All_without_IEA”; “use GO Term fusion” function was enabled; and only pathways with *P*<0.05 were shown. Full gene lists are available in supplementary table 1.

We used the STROBE cohort checklist when writing our report^28^.

## Results

The demographic, clinical and laboratory parameters of the volunteers included in the trial were previously published^18^. Briefly, 7 men and 2 women, completed the trial protocol. Their mean age was 45 (±9), 8 self-identified as Black, mean urine volume of all participants in a-24-hour collection was 3371 ml (Std±1599) on the first day of transition from American style diet to DASH diet. Selected parameters of our participants are presented in Table 1.

**Table 1:**
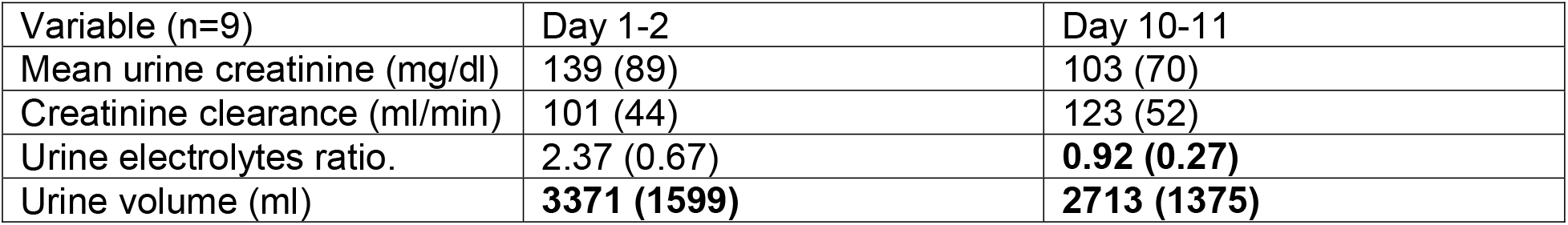
Selected parameters describing the kidney function changes of study volunteers between Days 1 and 11 of the trial. On Day 1-2 and 10-11 we performed a 24-hour urine collections, and the data for creatinine clearance and urine volume relate to that time period, whereas mean urine creatinine and urine electrolytes ratio relate only to days 1, 10. Data are shown as means (standard deviation).

| Variable (n=9) | Day 1-2 | Day 10-11 |
| --- | --- | --- |
| Mean urine creatinine (mg/dl) | 139 (89) | 103 (70) |
| Creatinine clearance (ml/min) | 101 (44) | 123 (52) |
| Urine electrolytes ratio. | 2.37 (0.67) | **0.92 (0.27)** |
| Urine volume (ml) | **3371 (1599)** | **2713 (1375)** |

### Separation and characterization of uEV’s from urine

The presence of uEVs was demonstrated by morphological (cryo-TEM) and molecular characterization (western blot analysis). The isolated uEV’s exhibited a spherical shape of various sizes and densities (Figure 2A,2B). Particle counting with NTA using Zetaview by Particle Metrix showed a median EV size of 166.1(84.0-323.3 nm). fNTA (fluorescence nanoparticle tracking) analysis of liposomes control showed 96% labeling, and our samples showed good labeling degree, ranging from 40 to 98% compared to the particles detected by light scatter (Supplementary table 6). There was no change in particle size distribution between day 1 and day 15 according to the scatter. The absence of Uromodulin (THP) indicates its successful removal by low ionic strength buffer (Figure 2C) from P20 and fractions 1+2 by SEC from P100 (Figure 2E). The presence of TSG101 associated with the exosome identity was confirmed by western blot analysis (Figure 2 D,F).

### Mass spectrometry analysis of uEV’s fractionation from the proof of concept study

a total of 1,593 proteins were identified in the combined P20+P100 fraction. The P20 fraction contained 1434 proteins with 34.4% identity when using 3 repetitions and 90.8 % identity between replicates when using 2 repetitions. The outlier specimen was attributed to technical challenges and was removed from further analysis and calculations. The P100 fraction contained 1229 proteins with 61.9 % identity between replicates (Supplementary Figure 2). 398 proteins were uniquely identified in P100 and 224 were uniquely identified in P20 (Supplementary Table 2). Figure 3 illustrates protein abundance patterns across the two separate fractions of P20 and P100, and their combination, showing that the combined sample looks like an average of the two individual fractions. There was a unique set of proteins identified in the combined cluster that did not cross the significance threshold of identification in either fraction alone (VPS28 Subunit Of ESCRT-I, UBE2N, CHMP4B).

**Figure 3:**
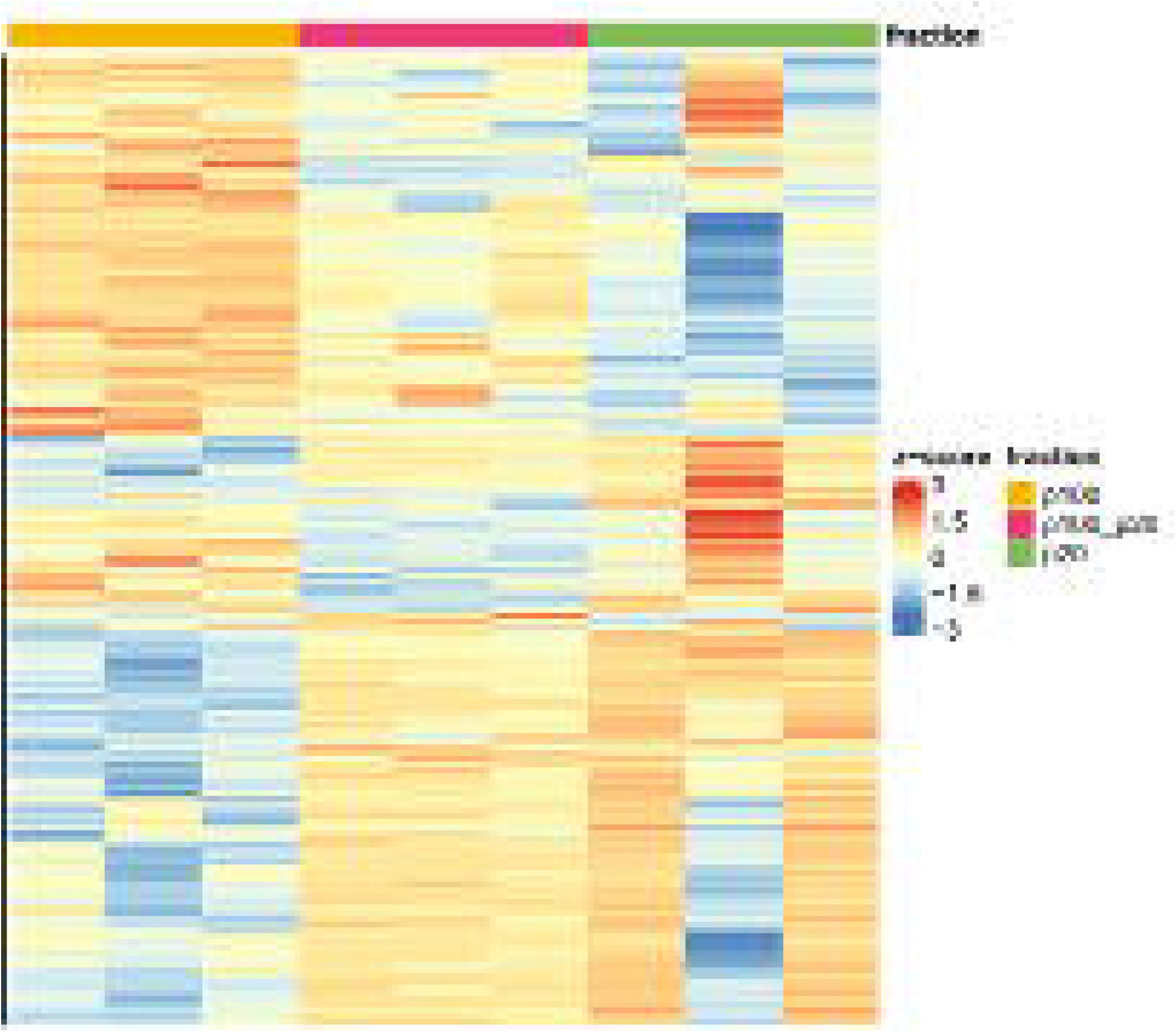
Heatmap showing protein expression patterns of exosomes from different uEV fractions. X axis shows the different fractions, left to right: P100, P100+P20, and P20. Y axis shows different proteins. Color scale relates to z-score where orange shades show increased expression and blue shades show decreased expression. The second column from the right, meaning the middle P20 sample, is the outlier that was removed from calculations

According to the DAVID annotation tool, both fractions were enriched for exosome proteins as 214 proteins from P20 were involved in this pathway (P-value of 7.0E-51) and 389 proteins from P100 were identified in this pathway (P-value of 7.0E-51). Proteins unique to P100 were affluent in pathways of exosome biosynthesis, multivesicular body sorting pathway, multivesicular body assembly and organization, and endosome transport. Full list of pathways can be found in Supplementary Figure 4.

Since both fractions were appropriately enriched for exosome terms, yet still had some differences in protein abundance, we chose to use the combined fractions and avoid information loss. The full list of proteins in fractions P20 and P100 can be found in Supplementary Table 3.

### Mass spectrometry analysis of the DASH intervention study

In the pooled P20 + P100 fractions we identified 1800 proteins across all 27 samples, 252 of them crossed the significance threshold of change (p-value < 0.05) at either day 5 or 11.

Between day 0 and 11 the expression of 65 proteins increased in a significant manner, including members of cytoskeleton organization and regulation, and protein complexes assembly. Expression of 77 proteins decreased including proteins from epithelial and neutrophil migration pathways. Full lists of proteins are available in Supplementary Table 5.

To focus our study on the proteins with consistent changes across the time course, we looked at those that change in both days 5 and 11 compared to day 0. We found that 22 proteins were upregulated and 25 were down regulated, and both sets of proteins were enriched for extracellular Exosomes pathway (p-value= 3.8E-14) including SLC12A3 (NCC) with higher abundance at both time points and Aquaporine 2 (AQP2) with reduced abundance. GO pathway analysis using DAVID showed that both the upregulated and downregulated protein lists were enriched for the extracellular exosomes pathway (p-value= 3.8E-14). AQP2 went down by 2.42 fold at day 5 compared to day 0 and down by 2.46 fold at day 11 compared to day 0 (Figure 4D) while NCC went up by 2.53 fold at day 5 compared to day 0 and up by 2.41 fold at day 11 compared to day 0 (Figure 4C). Full list of proteins, including fold changes in abundance, and GO pathway lists are available in Figure 4 and Supplementary Table 4. We explored expression patterns of proteins characteristic of the proximal nephron, to evaluate its involvement in the clinical changes we witnessed (e.g. AQP1, SGLT2, DPP4, Neprilysin, CD9, podocalyxin, Complement receptor type 1). These can be found in Supplementary Table 6. None of these proteins crossed the threshold for significant change of expression following DASH diet implementation during trial days.

**Figure 4:**
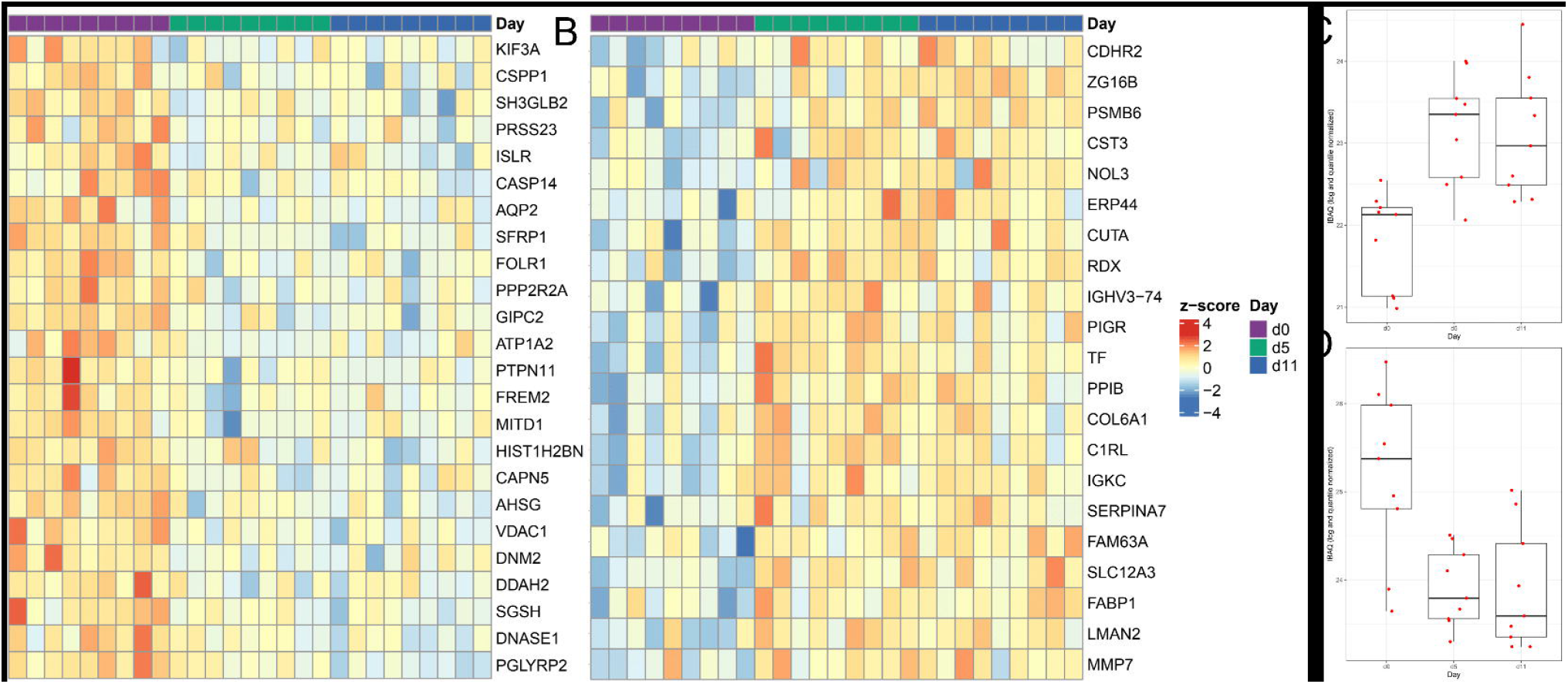
Heatmap of proteins with significant expression change over time. Panel A shows proteins with reduced expression, panel B shows proteins with increased expression. X axis shows all patients in the same order across the different days. Y axis shows the different clusters and names of the proteins. Color scale relates to z-score where orange shades show increased expression and blue shades show decreased expression. Panel C shows mean expression of NCC and Panel D mean expression of AQP2, according to iBAQ, across trial days.

## Discussion

Using a novel approach, we documented kidney epithelium protein abundance pattern changes in uEV in response to nutritional changes (Figure 1). Transitioning from American style diet to DASH diet challenges the kidneys to reverse the ratio of urine electrolytes – from high sodium and low potassium to low sodium and high potassium. This nutritional transition necessitates a change in abundance of channels in the kidney epithelium to discard the excess electrolytes. We characterized the change in channel abundance using uEV’s as a liquid biopsy, mirroring the alterations in the tissue. Proteomic analysis of uEV’s revealed an increase in expression of NCC and decrease in AQP2. Focusing on the preliminary protein changes following DASH diet implementation, this mechanistic trial potentially reveals the mechanism by which DASH diet decreases blood pressure.

Most of the uEVs proteome analyses published focus on the fraction we termed P100 and discard the low CF pellet (we termed P20). More recently, alternative methods to enrich uEV without carry over of Uromodulin (THP) for MS analysis have been proposed^29,30^ involving multiple techniques, often labor intensive with long centrifugation time for density gradient separations. Our group demonstrated recently that uEV separation from cell free urine with the low CF speed (e.g. P20) reveals a valuable and neglected source for uEVs^30^. In this study we now show that uromodulin can be easily separated from this P20 uEV pellet by simply washing the pellet with a low ionic strength buffer without the use of any reducing salt. This approach offers a uEVs fraction without uromodulin interference for an effective characterization of its proteome common to other studies [20]. However, to assure a comprehensive analysis in this study this P20 pellet was compared to the high CF P100 pellet, which is most often studied. Our pilot work demonstrates that simple washing with low ionic strength buffer was not sufficient to remove uromodulin from the P100 pellet, however depolymerization with 6M guanidine in acid condition followed by size exclusion chromatography successfully led to removal of uromodulin in the P100 pellet (Figure 2). Subsequent mass spectrometry showed that the proteome in P20 and P100 pellets (cleaned of uromodulin) are similar, but also contain specific clusters of proteins for each method (Figure 3). Hence, combining both P20 and P100 in one fraction offers a better coverage of the uEVs proteome and improves the sensitivity of detection and yield of the protein identification with more consistency across the triplicates of the pilot study. Similarity between P20 and P100 was confirmed with repeated EV extractions from the same sample, between pellets, and translated into similar pathways according to GO ontology using the DAVID annotation tool. We therefor conclude that our approach increases the identification of smaller EV’s that tend to cluster or co-sediment with uromodulin and increases detection yield, allowing us to identify key changes in protein abundance following DASH implementation, including decreases in AQP2.

The concept of using uEVs as a liquid biopsy to mirror tissue status in health and disease is contraversial. It is likely that the utility of uEVs depends on the context in which they are used. A previous report found no correlation between the level of tubular transport proteins in kidneys which had been removed for treatment of malignancy compared to uEVs collected before the extirpation^31^. It seems the absence of correlation is caused by the lag between uEVs and kidney tissue collection, which may have changed protein expression.

Other investigators found no difference in NKCC2 and NCC uEV excretion between salt-sensitive, salt-resistant, or salt-indeterminate hypertensive patients^32^. Yet the authors do not provide critical information regarding medication use, weight and height of the volunteers, or dietary intake, which could affect protein expression and the response to nutritional changes. Comparison was made between patients consuming different salt regimens rather than comparing the patients to themselves before and after a dietary intervention. This approach might have revealed the expected differences in uEV protein content that are salt dependent. However, a very recent study found that at a proteome level, independent of dietary conditions, a significant positive correlation exists between absolute protein abundances in uEVs and their abundance in the kidney^15^. This is reinforced by the strong correlation of the abundances of established uEV marker proteins (Alix, Tsg101, Cd63, and Cd81) in uEV pool and the kidney. The strongest significant correlation was for proteins classified as apical membrane transporters/channels^15^.

In our study, proteomic analysis of uEV’s revealed an increase in expression of NCC and decrease in AQP2 following DASH diet implementation. In addition to NCC and AQP2 other essential hormones such as Vasopressin (AVP), and aldosterone regulate body fluid homeostasis. For example, endogenous expression of AQP2 in mouse cortical CD principal cells (mpkCCDC14) is dramatically increased by administration of physiological concentrations of AVP^33^ while the major physiological role of aldosterone is to promote Na+ reabsorption^34^. Aldosterone may also influence cortical collecting duct (CCD) water permeability by changing intracellular AQP2 targeting and/or AQP2 abundance, in synergy with vasopressin. Aldosterone infusion in normal rats was associated with polyuria and decreased urine concentration^35^, and accompanied by decreased apical AQP2 labeling intensity in the connecting tubule and CCD^36^. Aldosterone reduced AQP2 mRNA and protein expression when administered together with AVP for short periods of time^37^. These data suggest that aldosterone tightly modulates AQP2 protein expression^33^. We saw an increase in aldosterone after DASH implementation in our study, which could explain the decrease in AQP2, and reduced water reabsorption, leads to increased urine volume, and eventually blood pressure reduction.

Also of note, the distal nephron has various response pathways to different physiological stimuli. In response to hypovolemia both ANGII and aldosterone are secreted to retain salt with minimal K+ secretion. This requires not only the presence of NCC, it must also be switched on by phosphorylation^38,39^. NCC activation in the DCT during low potassium intake causes an increase in fractional reabsorption of sodium. This reduces the electrochemical driving force for K+ secretion into the tubular fluid and potassium is conserved by reducing its excretion. Therefore, responses to maintain K+ homeostasis while consuming a low-K+/high-Na+ American style diet can raise blood pressure^40^. High potassium intake on the other hand leads to increase of aldosterone alone, and potassium is secreted without salt reabsorption^41^. Potassium secretion requires the excretion of sodium by reducing NCC activity (i.e dephosphorylation) and urinary Na+ loss increase analogous to a diuretic^42^.

When NCC is increased in uEVs following transition to DASH, sodium reabsorption is decreased and blood pressure is reduced? Protein abundance of NCC in the kidney and uEVs is generally the same^15^. The abundance of NCC in both kidney and uEVs is two-fold higher in rats on a low sodium than on a high sodium diet^32^. In uEVs isolated from mice fed a high potassium diet, protein levels of NCC, were significantly higher relative to controls^43^. In order to measure parallel protein abundance between uEV’s and kidney tissue in human subjects, trials were conducted in patients carrying known mutations in tubular transport proteins, because their genotype predicts protein abundance. Indeed, in patients with Gitelman syndrome NCC abundance is higher in uEVs, whereas in Gordon Syndrome it is lower compared to healthy subjects^44^. Following thiazide treatment, which blocks NCC, its abundance is increased in the rodent kidney^45^. This is also the case with essential hypertensive patients treated with thiazide^46^ diuretics. The increased abundance of NCC in uEVs, in our study, mirroring the tissue protein content, may be a compensatory effect to counteract reduced NCC activity due to inhibition.

This led us to reflect on the more general topic of whether uEV protein abundance mirrors the parental cell, a prerequisite to use uEV’s as a liquid biopsy. Changes in kidney protein abundance occur rapidly and constantly, leading to the high inter-subject variability in uEV protein abundance. To interrogate the differences between human subjects we should look for a more generalized and prolonged change in protein abundance that is reflected in uEVs. Such a stimuli could be compared in healthy vs. sick individuals^47^, or within the same person reacting to nutritional intervention^6^ or medication use^48^ or both. The response to these interventions may be captured more easily by uEV protein abundance.

DASH diet is a recommended lifestyle modification to reduce blood pressure^19^, but the actual mechanism is unknown. Our trial is the first to show the entire cascade of events from transition to DASH diet, to serum aldosterone increase, followed by change in protein abundance pattern in uEV and reversal of the urine electrolytes ratio (Figure 1). Initial studies of the DASH diet focused on electrolyte changes after 2 or 4 weeks. This analysis dissects the response to nutritional changes in the immediate period of changes and at a resolution of the ion channel level by use of uEV’s, as a surrogate to invasive procedure. Our results suggest that DASH diet related blood pressure reduction is the result of reduced water and sodium reabsorption, potentially through changes in availability of AQP2 and NCC.

Decreases in AQP2 increases urine volume, reduces serum/blood volume and subsequently reduces blood pressure. The increase in NCC implies an attempt to compensate for inhibition of the ion channel, similar to thiazide treatment, even though no exogenous substance is used. This inhibition results in an increase of the electrical driving force for potassium secretion, which is swiped with the sodium.

Our study has some limitations. First, the study has a small sample size (n=9). This was a proof-of-concept trial which implemented a two-week inpatient protocol. Moreover, the inclusion criteria narrowed the study population, to exclude physiologic biases such as use of anti-hypertensive medication and obesity, so the generalizability of our findings to those populations is not yet known. Despite the small cohort, we were able to reach statistical significance in protein abundance patterns following the intervention. Another limitation is the lack of phosphorylation information of the different ion channels.

Nutritional changes are a very subtle intervention and initiate a cascade of events leading to differences in availability of ion channels, dictated by phosphorylation. Future studies could focus on differences in phosphorylation status of uEVs proteins by mass spectrometry and precipitation methods using specific antibodies. Our uEVs extraction protocol is improved compared to previous approaches, but still would benefit from further refinement. We used guanidine, which can damage EV cargo and increase polydispersity, and the column we used for SEC is not sensitive enough to identify the small EVs, which are lost because of co-sedimentation with THP. These technical issues should be approached in future studies aiming at increased yield of EV’s extraction.

Our study has some important strengths. This meticulously planned protocol implemented nutritional modifications on human subjects, representing physiological changes. Use of human subjects to investigate proteomic response has been limited by the need of invasive procedures such as kidney biopsy. This led to the use of animal models and cell cultures that have their limitations. The innovative use of uEVs as a non-invasive easy to acquire surrogate marker for an invasive procedure has enabled insight into the tissue at the protein abundance level. It’s use can likely to be expanded to investigate kidney responses in other settings.

Another strength is the time-dependent changes we describe. This is the first-time protein abundance changes are captured by uEVs simultaneously with nutritional changes, indicating causality. Previous studies have focused on the effectiveness of DASH as blood pressure reducing tool a few weeks after nutritional transition thereby missing the ion channel modulation enabling the clinical endpoint.

## Conclusion

Nutrition is an essential tool in reducing hypertension, a modifiable cardiovascular risk factor and DASH diet is a proven and recommended intervention to reduce blood pressure. Our study has provided insights into a potential mechanism by which DASH diet lowers blood pressure by increasing urine volume and decreasing sodium reabsorption. These clinical changes coincide with a change in protein abundance patterns, mainly an increase in NCC and decrease in AQP2 which account for the sodium excretion and handling free water. Future studies should focus on the interplay between vasopressin and aldosterone as determinants of AQP2 expression, and on aldosterone as a blood pressure reducing hormone, exonerating it from being the eternal cardiovascular villain.

## Data Availability

All data produced in the present study are available upon reasonable request to the authors

## Abbreviations

(DASH): Dietary Approach to Stop Hypertension
(BP): blood pressure
(uEV): urinary extracellular vesicles
(AQP2): Aquaporin 2
(NCC): sodium potassium symporter

## Author contribution

DB- Conceptualization, Data curation, Investigation, Methodology, Visualization, Writing – original draft, Writing – review & editing, LM –exosome extraction, Data curation, Investigation, Software, Validation, Writing – review & editing HM – proteomic analysis, Data curation, Software, Validation, Writing – review & editing DB – bioinformatic analysis, Data curation, Formal analysis, Software, Validation, Writing – review & editing TC – bioinformatic analysis, JNT – supervision, writing – review & editing, RGK – supervision, writing – review & editing, UE – supervision, methodology, editing.

## PX Complete

The mass spectrometry proteomics data have been deposited to the ProteomeXchange Consortium via the PRIDE [1] partner repository with the dataset identifier PXD036820.

## Reviewer account details

Username: Password: LVO6IKFZ

## Trial registration

clinicaltrials.gov ID: NCT04142138

