## supplementary material for "Urinary Extracellular vesicles abundance of SLC12A3 (NCC) increase and Aquaporine2 decrease following DASH diet implementation"

Supplementary materials and methods:

Supplementary Tables 1,2,3 appear in the attached excel spreadsheet.

Supplementary table 1: proteins uniquely expressed in either P20 or P100.

Supplementary table 2:Proteins identified in either fraction P20 or P10 or a combination of the two.

Supplementary table 3: Proteins with expression change from day 0 to day 5, from day 0 to day 11, and from day 0 to both day 5 and 11.

Supplementary table 4: Proteins with expression change from day 0 to days 5 and 11. Lists show protein names

| Day to day 5 and day 11 | |
| --- | --- |
| Upregulated | Down regulated |
| Zymogen granule protein 16 homolog B  Proteasome subunit beta type-6;Proteasome subunit beta type  Solute carrier family 12 member 3  Protein FAM63A  Protein CutA  Serotransferrin  Vesicular integral-membrane protein VIP36  Endoplasmic reticulum resident protein 44  Fatty acid-binding protein, liver  Nucleolar protein 3  Cadherin-related family member 2  Peptidyl-prolyl cis-trans isomerase B  Thyroxine-binding globulin  Cystatin-C  Matrilysin  Ig kappa chain C region  Polymeric immunoglobulin receptor;Secretory component  Collagen alpha-1(VI) chain  Complement C1r subcomponent-like protein  Radixin | N-acetylmuramoyl-L-alanine amidase  Secreted frizzled-related protein 1  Histone H2B;Histone H2B type 1-L;Histone H2B type 1-M;Histone H2B type 1-N;Histone H2B type 1-H;Histone H2B type 2-F;Histone H2B type 1-C/E/F/G/I;Histone H2B type 1-D;Histone H2B type F-S;Histone H2B type 1-K;Histone H2B type 1-A  N-sulphoglucosamine sulphohydrolase  Alpha-2-HS-glycoprotein;Alpha-2-HS-glycoprotein chain A;Alpha-2-HS-glycoprotein chain B  PDZ domain-containing protein GIPC2  Deoxyribonuclease-1  Folate receptor alpha  Aquaporin-2  Endophilin-B2  Tyrosine-protein phosphatase non-receptor type 11  Serine/threonine-protein phosphatase 2A 55 kDa regulatory subunit B alpha isoform  Centrosome and spindle pole-associated protein 1  Caspase-14;Caspase-14 subunit p17, mature form;Caspase-14 subunit p10, mature form;Caspase-14 subunit p20, intermediate form;Caspase-14 subunit p8, intermediate form  FRAS1-related extracellular matrix protein 2  Voltage-dependent anion-selective channel protein 1  Serine protease 23  MIT domain-containing protein 1  Immunoglobulin superfamily containing leucine-rich repeat protein  Kinesin-like protein;Kinesin-like protein KIF3A  N(G),N(G)-dimethylarginine dimethylaminohydrolase 2  Sodium/potassium-transporting ATPase subunit alpha-2  Dynamin-2  Calpain-5 |

Supplementary table 5: NTA from previous exosome extraction. Urine samples from participants 3,6,7,11,12 taken on days 1 and 15 of the trial were analyzed. They were compared to pooled first void urine from 6 healthy individuals, a positive control of exosomes and a negative control of liposomes.

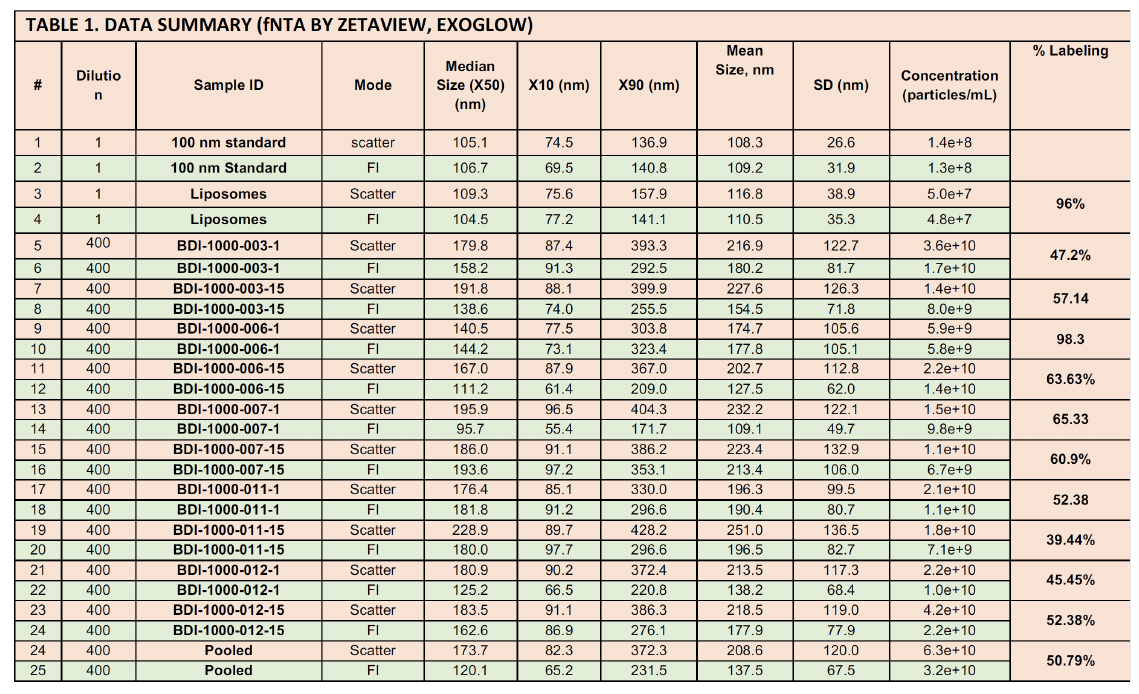

Supplementary figure 1: Venn diagram visualizing the resemblance and difference of protein content between repetitions of uEV’s extraction from the same sample. One healthy volunteer gave a first void urine sample that was processed for exosomes and proteomics in three independent repetitions. Panel A – P20, 1434 proteins were identified, one repetition was technically unsuccessful, and including it reduced the similarity between samples to 34%. When excluding this sample, identity between the remaining two samples increased to 90.8%. Panel B – P100, 1292 proteins were identified, identity between repetitions was 61.9%. Panel C – a combination of P20 and P100. 1561 proteins were identified. Identity of the three repetitions was 77.6%.

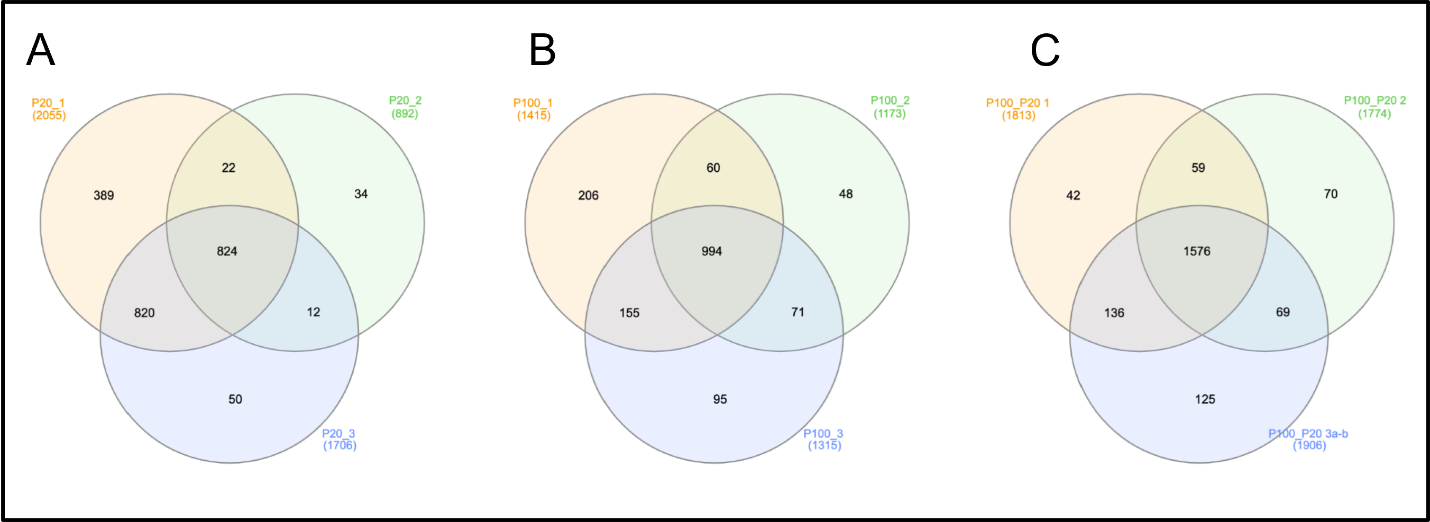

Supplementary figure 2: Dissolving THP from P100. Silver staining of size exclusion chromatography fractionation of P100. Panel A – no reducing agent used, THP appears in the first three fractions. Panel B – using TCEP in PH=7.8 THP still appears across all fractions. Panel C – using Guanidine and Urea PH=3.5 reduces THP from fractions 1 and 2.

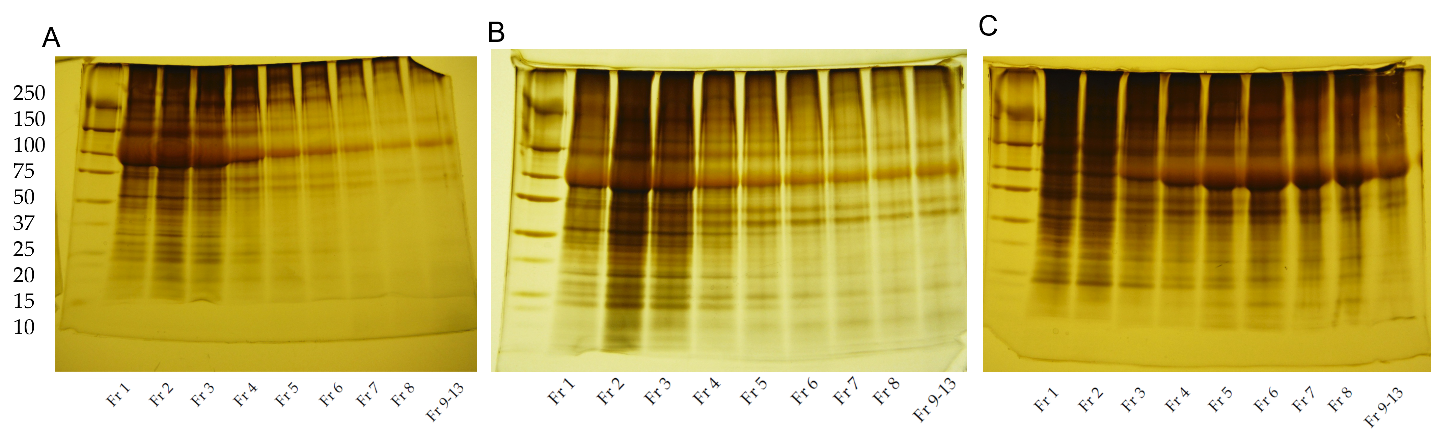

Y axis – size ladder, x axis – different fractions eluted by SEC.

Supplementary figure 3: functional pathways identified by DAVID annotation tool. Proteins uniquely expressed in either fraction P20, P100 or the combination were uploaded to the annotation tool. The Y axis lists the pathways identified according to the number of proteins involved in descending order. X axis shows log (base 10) of the pvalue, multiplied by -1.

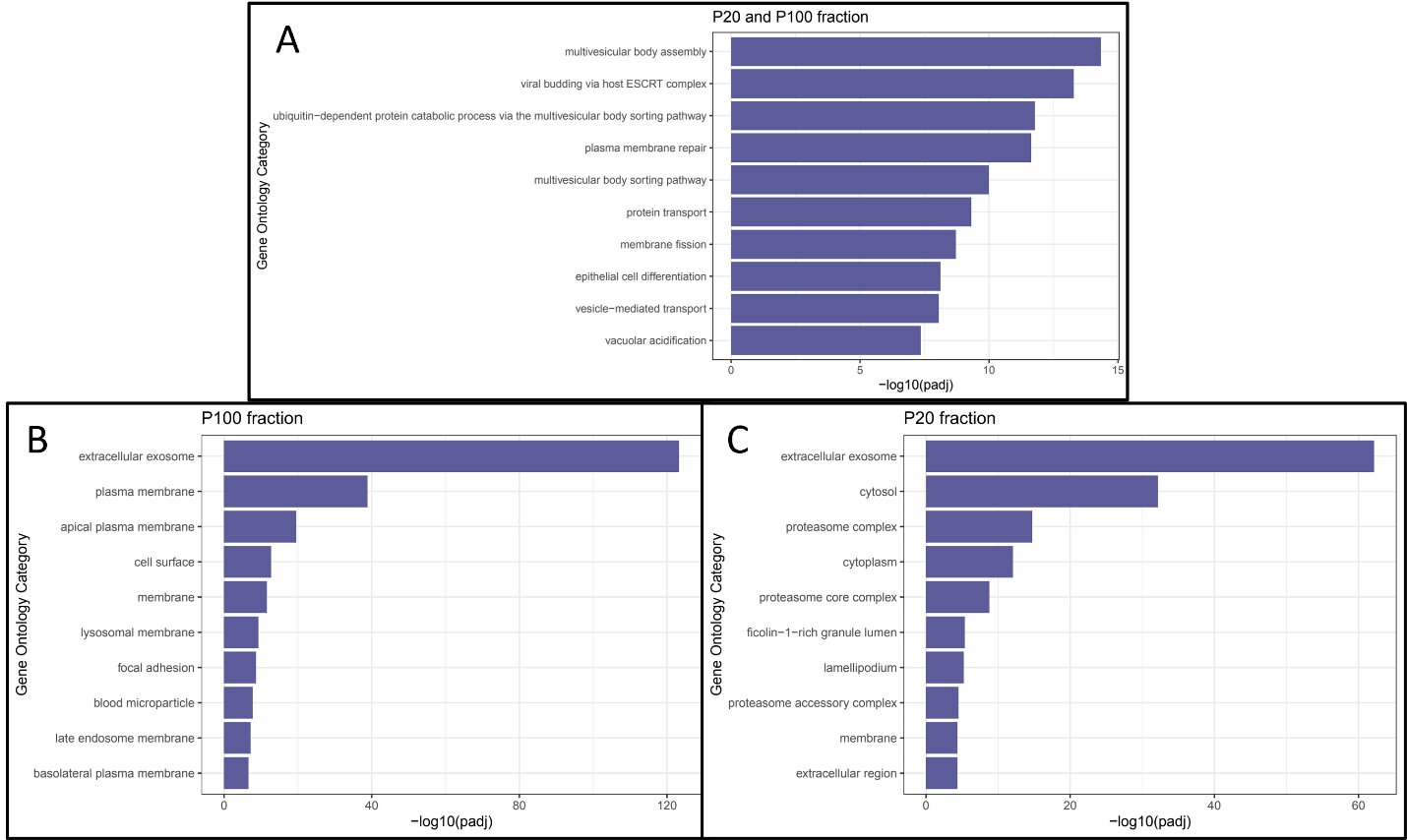

**Supplementary methods description**

Majority of the chemical reagents were purchased from Sigma-Aldrich unless otherwise specified.

**Urinary Extracellular Vesicle enrichment.**

For the pilot study urine was centrifuged at a Relative Centrifugal Force (RCF) of 4,600g at max radius 168 mm (5000 rpm) in a TX-400 Sorvall ST16R (Thermo Fisher Scientific, Waltham, MA)) swing bucket rotor (k Factor 9153) for 30 minutes at room temperature (RT) (braking set at 9). Fifteen millilitres of supernatant 4,600g (SN4,600) for the pilot study and SN from the Dash diet cohort were centrifuged at 16,000 rpm for 30 minutes at 4°C using the Sorvall RC 5B centrifuge (Sorvall) and the SS34 rotor (k-factor 714 at maximum speed). Beckman polycarbonate bottle 50 mL (Dimensions: 29 X 104 mm cat number 357002 ) were filled with 15 ml of urine. The Relative centrifugation force (RCF) span approximately from 20,000 g (average radius) to 32,000g (max radius). We refer to this uEVs pellet as **P20**. P20 was solubilized with 1 mL of low ionic strength buffer made of 10 mM HEPES pH7.4 + 2.5 mM EDTA pH 8.0 transferred to 1.5 mL microcentrifuge tubes (Axygen; Corning Inc. Corning, NY) and centrifuged at 15,000 rpm; RCF 21,130 g) for 30 minutes at 4°C in an Eppendorf microcentrifuge 5424 fix angle rotor (FA-45-24-11) (Eppendorf, Hamburg,Germany). This step was repeated twice for a total of 3 solubilization-centrifugation cycles. Pellets were stored at -80°C degrees as pellet and solubilized after thawing in 0.1 μm filtered (Minisart^®^ PES syringe filter code 16553-------K, Sartorious, Göttingen, Germany) phosphate buffered saline solution (PBS) pH 7.4 no Ca^2+^ and Mg^2+^ (Gibco^®^, life technology™, Thermo Fisher Scientific) (PBS**^0.1μm^**) or electrophoresis solubilisation buffer (ESB).

The urine supernatant of P20 (SN20) was further centrifuged in Beckman Coulter XL-80 ultracentrifuge using the 70Ti fix angle rotor (k-factor 44 at maximum speed) (Beckman) for 2 hours at 40,000 rpm at 4°C, using polycarbonate tubes (Beckman catalogue number 35563). RCF span between 164,000 g (max radius) to 117,000 g (average radius). We refer to this uEVs pellet as **P100.**

**Depletion of Tamm-Horsfall Protein from uEV P100 by size exclusion chromatography (SEC)**

P100 pellets were solubilized in 200 μL of a solution made of 6 M guanidine HCl, 0.1 m citrate buffer pH3.5; 50 mM arginine and 50 mM 6-amicaproic. Final pH 3.5 to depolymerize and unfold THP filaments. Separtaion of EVs from THP was performed using a qEV70 original column (Izon) connected to automatic fraction colletor (AFC) unit (Izon). Two mL of guanidine solution was flowed in the column before loading P100 solubilize in the guanidine solution for 20 minutes in an end-over-end rotation at RT. After loading P100 1 mL of guanidine solution was added followed by a mobile phase made of : 10 mM HEPES pH 7.4 + 2.5 mM EDTA + 0.1 % sodium azide filtered with 0.1 μm syringe filter (Minisart^®^ PES syringe filter code 16553K, Sartorious, Göttingen, Germany). One mL fractions were collected.

### The protein content was precipitated by 20 % (v/v) trichloracetic acid (TCA) supplemented with 0.08 % (w/v) sodium deoxycholate (DOC)(1). Briefly, 250 μL of 100 % (v/v) TCA + 0.4 % (w/v) DOC was added to 1.0 mL of the chromatography fractions, vortexed and incubated in ice for 30 minutes. Samples were then centrifuged at max speed (15,000 rpm; RCF 21,130 g) for 30 minutes at 1°C using the Eppendorf microcentrifuge 5424 fix angle rotor (FA-45-24-11). Supernatant was discarded and the pellet was added 800 μL 100 % (v/v) acetone and incubated at -20°C overnight and then centrifuged max speed (15,000 rpm; RCF 21,130 g) for 30 minutes at 1°C in the microcentrifuge. Supernatants were discarded and the pellets dried in the fume hood for 10 minutes to be finally solubilized in either ESB or of PBS0.1μm respectively and transferred in a 1.5 mL Posi-Lock™ Low-Binding Microcentrifuge Tubes (Thomas Scientific; Catalogue number 1149X75)

**Lipid extraction for MS analysis**

P20 pellets and P100 TCA-DOC pellets were resolubilize in 100 μL of PBS**^0.1μm^** and delipidated by chloroform methanol(2). Briefly, 400 μL of 100 % (v/v) methanol was added to the sample, vortexed and centrifuged for 10 seconds at 9000g. Two hundred μL of chloroform were added, vigorously vortexed and centrifuged for 10 seconds at 9000g. Three hundred μL of deionized water was added, mixed vigorously and centrifuged for 5 minutes at 9000g. The aqueous upper phase was discarded and the interface protein layer was precipitated by adding 300 μL of 100 % (v/v) methanol and centrifuged for 10 minutes at max speed.

**Protein assay, Gel electrophoresis and Western blot**

uEVs and protein pellets were solubilised in 40 μL of ESB made of: 6 M urea (Bio-Rad Laboratories, Hercules, CA), 2 M thiourea, 5% (w/v) sodium dodecyl sulphate (SDS) (Bio-Rad Laboratories), 40 mM Tris-HCl, pH 6.8, 0.5 mm ethylenediaminetetraacetic acid (EDTA) (Bio-Rad Laboratories), 20% (v/v) glycerol and 50 mM dithiothreitol (DTT) (Bio-Rad Laboratories) (3). Samples were denaturized overnight at room temperature (RT). Proteins were separated by hand cast SDS-PAGE gradient gels (Resolving gel T= 5-20 % (w/v); C=2.6 %; Stacking gel T= 3.5 % (w/v); C=2.6 %) in 25 mM Tris (Bio-Rad Laboratories), 192 mM glycine (Bio-Rad Laboratories) and 0.1 % (w/v) SDS (Bio-Rad Laboratories) buffer (4) and either stained with colloidal Coomassie G-250(5)or silver nitrate(6) or transferred onto a 0.45μm nitrocellulose membrane (Amersham™ Protean™ 0.45μm NC, GE Healthcare) in a wet transfer system buffer made of 25 mM Tris, 192 mM glycine and 20 % (v/v) methanol for 2 hours at 200 mA per gel in ice bath(7). Nitrocellulose membranes were saturated with Odyssey blocking buffer (LI-COR Biosciences, Lincoln, NE) and incubated in: 0.5 μg/mL rabbit anti tumor susceptibility gene 101 protein (TSG101) (Sigma-Aldrich, catalogue number T5701); 1.0 μg/mL mouse anti ALIX (ThermoFisher Scientific clone 3A9 catalogue number MA1-83977); 1.0 μg/mL biotin mouse anti human CD9 (clone HI9a), 1.0 μg/mL biotin mouse anti human CD81(TAPA-1) (clone 5A6), biotin mouse anti human CD63 (clone H5C6), 1.0 μg/mL mouse anti human CD35 or complement receptor 1 (CR1) (clone 9H3) and 1.0 μg/mL	mouse anti human CD26 or dipeptidyl dipeptidase 4 (DPP4) (clone BA5b) (Biolegend catalogue numbers: 312112, 349514, 353018, 332402 and 302718 respectively) 1.0 μg/mL mouse anti Aquaporin-2 (AQP2, clone E2) and 1.0 μg/mL mouse anti sodium/glucose cotransporte-2 (SGLT2, Clone D-6)(Santa Cruz Biotechnology, catalogue number sc-515770 and sc-393350 respectively); 1.0 μg/mL	goat anti neprilysin/CD10 ( R&D System catalogue number AF1182); 1.0 μg/mL mouse anti podocalyxin (PODXL, clone 3D3) (Novus Biological catalogue number NBP2-25219) and 1.0 μg/mL	rabbit anti syntenin-1 (EPR8102) ( Abcam catalogue number ab133267) in the Odyssey blocking buffer diluted 1:1 with in house PBS (10 mM sodium phosphate dibasic, 1.8 mM potassium phosphate monobasic, 137 mM sodium chloride, 2.7 mM potassium chloride ) and 0.15 % (v/v) Tween-20. After 3x10 minute washes in PBS-Tween (0.15%, v/v), membranes were incubated with goat anti mouse (code 925-68070 and/or 925-32210), goat anti rabbit (code 925-68071 and/or 925-32211), streptavidin (code 925-68079 and/or 925-32230) and donkey anti goat (code 925-68074) either red (displayed in red colour excitation 680 nm, emission 700 nm) or infrared (displayed in green colour excitation 780 nm, emission 800 nm) dye-coupled secondary antibody 0.1 μg/mL (LI-COR Biosciences, Lincoln, Nebraska, USA) in an Odyssey blocking solution diluted at 1:1 with PBS and 0.15 % (v/v) Tween-20; 1hour at RT. Acquisition of the fluorescent signal was performed by Odyssey infrared imaging system with resolution set at 169µm (LI-COR Biosciences,).

**Nanoparticle Tracking Analysis (NTA)**

NTA was performed using the ZetaViewPMX 120 (Particle Metrix, Meerbusch, Germany) configured with a 488 nm laser with a long wave-pass (LWP) cut-off filter (500nm) and a sensitive CMOS camera 640 x 480 pixels. Each P21sample was diluted in 2 mL of 0.1 μm filtered (Minisart^®^ high flow hydrophilic 0.1 μm syringe filter Sartorious) deionized water (DI 18 MΩ/cm) [34] to obtain a particle concentration between 1 x 10^7^ and 1 x 10^8^ particles/mL. The instrument was set to a constant temperature of 25°C, a sensitivity of 70, a shutter speed of 80 and a frame rate of 30 frames per second (fps). Each sample was measured at 11 different positions throughout the cell, with 5-7 cycles of readings at each position in order to have a minimum of 1000 traces. Post-acquisition parameters were set to a minimum brightness of 30, a maximum size of 200 pixels, and a minimum size of 5 pixels. Automated cell quality control was checked using high quality deionized water (DI). Camera alignment and focus optimization was performed using polystyrene 100 nm beads (ThermoFisher Scientific catalogue number 3100A). Data analysis was performed with ZetaView 8.02.28 software provided by the manufacturer. Automated report of the particles recording across the 11 positions were manually checked and any outlier position was removed to calculate particle concentration and distribution expressed by mode, median and mean.

**Cryo-Transmission Electron Microscopy (Cryo-TEM)**

Low speed centrifuged uEV P21 and P21^TCEP^ pellets were solubilized in 20 μL PBS**^0.1μm^** and applied to a glow-discharged, perforated carbon-coated grid (2/2-3C C-Flat; Protochips, Raleigh, NC, USA), manually blotted with filter paper and rapidly dipped into liquid ethane. Grids were stored in liquid nitrogen, then transferred to a Gatan 626 cryo specimen holder (Gatan, Warrrendale, PA, USA) and kept at −180°C. Low-dose images were collected at a nominal magnification of 29,000× on the Tecnai F20 Twin transmission electron microscope operating at 120 kV. Digital micrographs were recorded on a Gatan US4000 charge-coupled device camera(8). Cryo-TEM was performed in the molecular electron microscopy core at the University of Virginia (<https://med.virginia.edu/molecular-electron-microscopy-core/services/>).
